# Excess Mortality in the Vaccination Era in the United States, By State and 6-Month Period

**DOI:** 10.1101/2023.03.07.23286927

**Authors:** Jeremy Samuel Faust, Benjamin Renton, Chengan Du, Alexander Junxiang Chen, Shu-Xia Li, Zhenqiu Lin, Harlan M. Krumholz

**Affiliations:** Brigham and Women’s Hospital, Harvard Medical School, Boston, Massachusetts; Ontos Analytics, Cambridge, Massachusetts; Center for Outcomes Research and Evaluation, Yale University School of Medicine, New Haven, Connecticut; Harvard University, Cambridge, Massachusetts

## Abstract

**Introduction:** The US continued to record all-cause excess mortality after the rollout of vaccines. We sought to quantify excess mortality by state and compare these rates to primary series vaccination completion levels.

**Methods:** Observational cohort, US and state-level data. Expected monthly deaths were modeled using pre-pandemic US and state-level data (2015-2020). Mortality data was accessed from CDC public reporting.

**Results:** We find that in a two-year period since the rollout of vaccines, the US recorded >874,000 excess deaths. Vaccination rates and excess mortality were most strongly correlated in first two periods before the Omicron variant.

**Conclusion:** The association between vaccination and lower excess mortality rates was strongest in 2021 and early 2022, prior to high population rates of infection-acquired immunity. The findings underscore the benefits of the rapid vaccination rollout campaign and the continued need to boost at-risk populations.

## Introduction

Vaccination has been shown to reduce COVID-19 mortality, the major driver of all-cause excess mortality.^1^ Both vaccination rates and excess mortality has varied across the US.^2^ We sought to determine the relationship of state-level excess deaths and vaccination rates across different waves of the pandemic. We hypothesized there would be a strong relationship between vaccination rates and excess mortality among US adults and this relationship would weaken as more people had been infected with SARS-CoV-2.

## Methods

As previously, we applied seasonal autoregressive integrated moving average (sARIMA) modeling to monthly all-cause death data from the Centers for Disease Control and Prevention and state population counts (2014-2020) to create monthly population and expected death estimates for the study period (January 2021-December 2022), correcting for population decreases owing to cumulative pandemic-associated excess deaths.^3,4^ Excess deaths were calculated (observed minus expected deaths).

The study was divided into four six-month periods: January 1-June 30, 2021 (Early Vaccine Rollout), July 1-December 31, 2021 (Delta), January 1-June 30, 2022 (Omicron 1), July 1-December 31, 2022 (Omicron 2). For all states, expected deaths for three age groups of adults were modeled (18-49, 50-64, and ≥65 years), which were then combined, creating a composite excess mortality figure. Vaccination rates were calculated as the primary series completion rate for each age group and state at the middle date of each period.^2^

Analyses were conducted with R version 4.0.3. This study used publicly available data and was not subject to institutional review approval per HHS regulation 45-CFR-46.101(c).

## Results

During the total two-year study period (vaccine era), the US recorded 874,688 (95% CI, 751,497-997,879) excess deaths. There were 219,239 (95% CI, 175,703-262,776) excess deaths during the Early Vaccine Rollout, 335,582 (95% CI, 291,680-379,484) excess deaths during the Delta, 196,788 (95% CI, 152,887-240,689) excess deaths during the Omicron 1 and 123,078 (95% CI, 80,063-166,094) excess deaths during Omicron the Omicron 2 periods, corresponding to 85, 130, 76 and 47 excess deaths per 100,000 persons, respectively. Vaccination rates and excess mortality were inversely correlated in the first three 6-month periods (January 2021-June 2022), with the strongest correlations observed during the Delta period (Figures 1 and 2). There was no observed correlation during the later Omicron period (July-December, 2022) (Figures 1 and 2). These findings were seen in all adults (Figure), including among adults ages ≥65.

**Figure 1.**
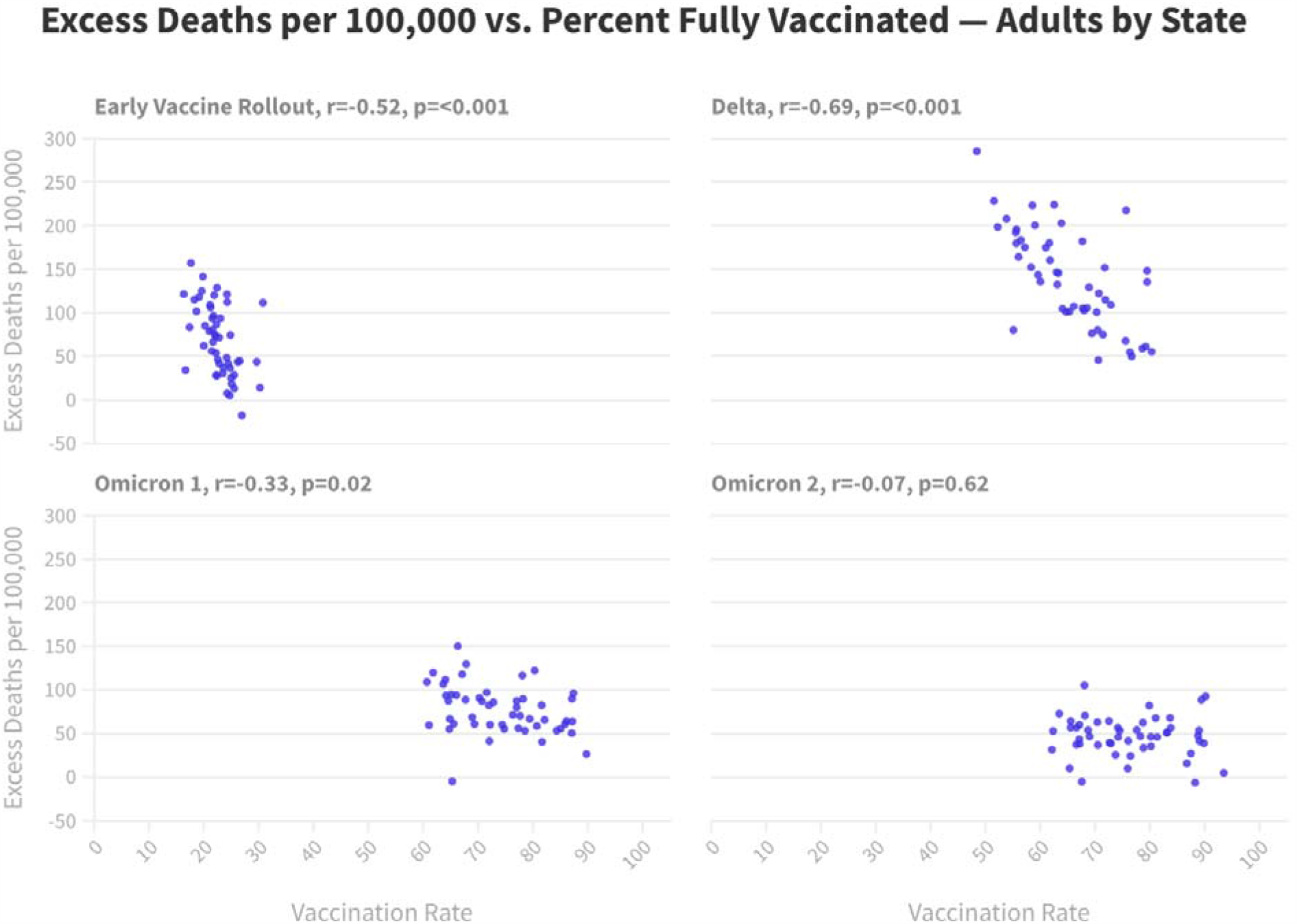

**Figure 2.**
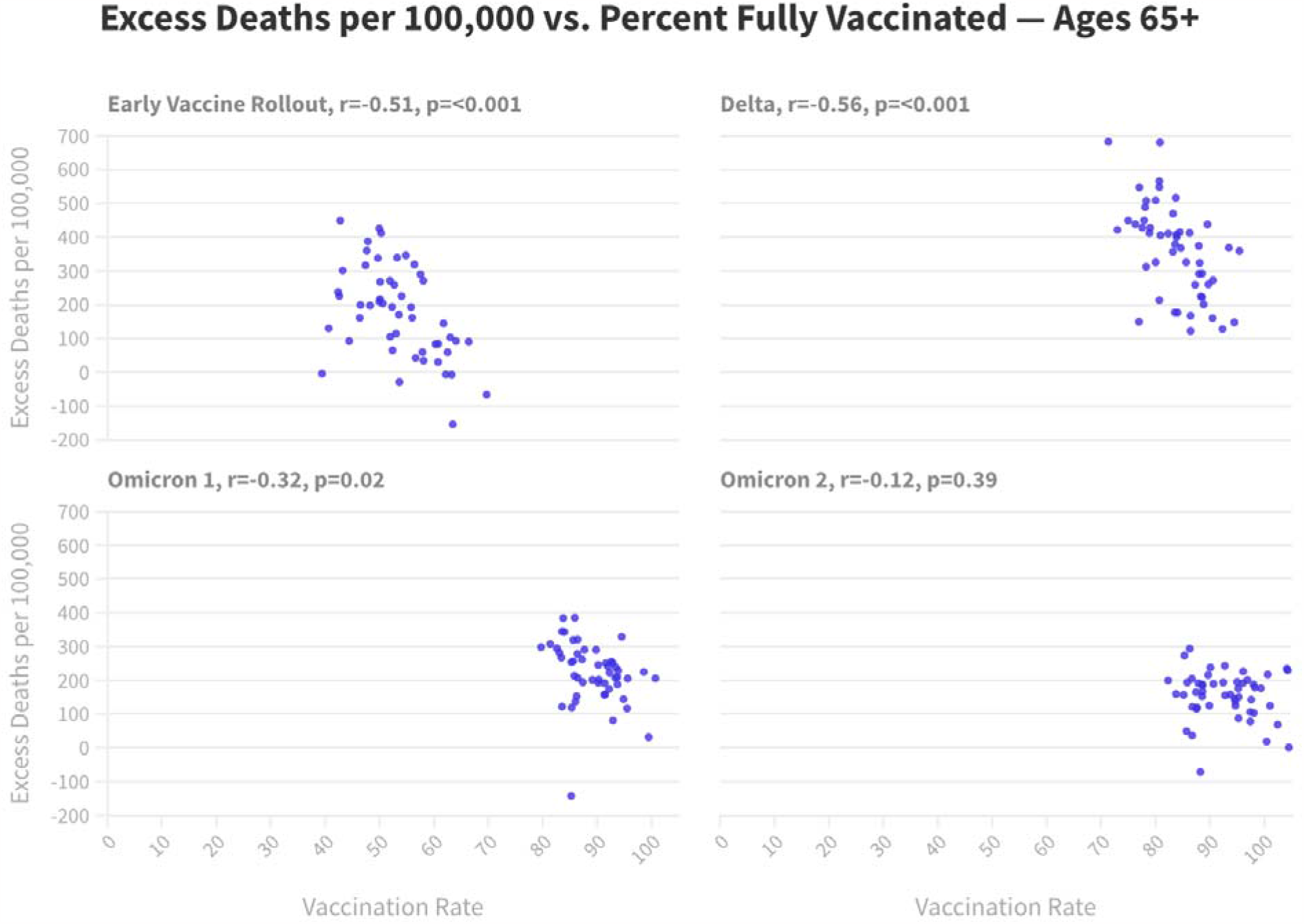

## Discussion

There were differences in excess mortality across US states during from January 2021-December 2022 which varied by vaccination rate. A stronger relationship was observed earlier in the vaccination period of the pandemic, when natural immunity was lowest. The loss of an association between excess mortality and vaccination rates later in the study (including in adults ages ≥65) argues for the necessity of boosting at-risk populations, given what is known about booster effectiveness.^5^ The results also imply that the campaign to rapidly vaccinate the population prior to high rates of immunity acquired by infection saved many lives. This study also suggests the potential lives that could have been saved if vaccination rates had been higher, were this relationship to be causal, a reasonable assumption based on the vaccine trials.^5,6^ Limitations include its observational nature and its use of provisional mortality data.

## Data Availability

All data produced in the present study are available upon reasonable request to the authors

## Author Contributions

Dr. Faust and Mr. Renton had full access to all of the data in the study and takes responsibility for the integrity of the data and the accuracy of the data analysis.

### Concept and design

Faust, Du, Krumholz, Renton. *Acquisition, analysis, or interpretation of data:* All authors. *Drafting of the manuscript:* Renton, Faust.

### Critical revision of the manuscript for important intellectual content

All authors.

### Statistical analysis

Faust, Du, Li, Lin, Renton. *Administrative, technical, or material support:* Faust, Du, Lin. *Supervision:* Faust, Krumholz.

### Data visualization

Renton.

